# Prevalence of Autoimmune Diseases in Functional Neurological Disorder: Influence of Psychiatric Comorbidities and Biological Sex

**DOI:** 10.1101/2023.10.13.23296897

**Authors:** Anna Joseph, Gaston Baslet, Mary A O’ Neal, Ginger R Polich, Irene Gonsalvez, Andrea N Christoforou, Barbara A Dworetzky, Primavera A Spagnolo

## Abstract

**Background:** Functional neurological disorder (FND) is a common and disabling neuropsychiatric condition, which disproportionally affects women compared to men. While the etiopathogenesis of this disorder remains elusive, immune dysregulation is emerging as one potential mechanism.

**Aim:** To begin to understand the role of immune dysfunction in FND, we assessed the prevalence of several common autoimmune diseases (ADs) in a large cohort of patients with FND and examined the influence of psychiatric comorbidities and biological sex.

**Methods:** Using a large biorepository database (Mass General Brigham Biobank), we obtained demographic and clinical data of a cohort of 643 patients diagnosed with FND between January 2015 and December 2021. The proportion of ADs was calculated overall, by sex and by presence of psychiatric comorbidities.

**Results:** The overall prevalence of ADs in our sample was 41.9%, with connective tissue and autoimmune endocrine diseases being the most commonly observed ADs. Among patients with FND and ADs, 27.7% had ≥2 ADs and 8% met criteria for multiple autoimmune syndrome. Rates of ADs were significantly higher in subjects with comorbid major depressive disorder and post-traumatic stress disorder (*p*=□0.02). Women represented the largest proportion of patients with concurrent ADs, both in the overall sample and in the subgroups of interest (*p*’s□<□0.05).

**Conclusions:** This study is unique in providing evidence of an association between FND and ADs. Future studies are needed to investigate the mechanisms underlying this association and to understand whether FND is characterized by distinct dysregulations in immune response.

## 1. Introduction

Functional neurological disorder (FND) is a complex and disabling neuropsychiatric condition presenting with a variety of sensorimotor symptoms and alterations in cognitive and emotion processing [1,2]. FND is also frequently associated with psychiatric comorbidities, including major depressive and anxiety disorders, and posttraumatic stress disorder (PTSD) [3], as well as with other neurological diseases [4,5]. With an annual incidence of 4–12 per 100□000, FND is a disabling disease which disproportionally affects women compared to men [6].

The diagnostic criteria and therapeutic approaches to this disorder are currently well-established, albeit diagnosing FND still poses several challenges and available treatments have shown limited efficacy [1,7]. Therefore, researchers in the field are devoting substantial efforts to uncovering the pathophysiological mechanisms underlying FND symptoms, with the goal of identifying disease biomarkers and novel therapeutic alternatives.

Recent findings suggest that FND is associated with neuroendocrine and immune abnormalities [8]. Specifically, patients with this disorder show blunted morning cortisol secretion [9–11], alterations in peripheral inflammatory markers, such as C-reactive protein, interferon-gamma, interleukin-6, and tumor necrosis factor-alpha [12–14], and presence of anti-ganglionic acetylcholine receptor antibodies associated with autonomic dysfunction [15,16]. Furthermore, a recent hypothesis-free genome-wide epigenetic study identified methylomic changes at the level of multiple immune-related pathways in individuals with FND with and without history of childhood abuse [17]. However, other studies have found no evidence of immune dysregulation in FND patients (for a review see [8]).

Discrepancies between such findings can be explained by several factors including study design and sample size. In addition, sex-related biological factors may be particularly relevant. It is well-established that immune responses and inflammation strongly differ between sexes, with females showing heightened proinflammatory activity compared to males [18]. Second, women have a two-fold higher risk to develop psychiatric disorders commonly comorbid to FND, such as major depressive disorder (MDD) and PTSD [19,20]. Such disorders have been associated with low-grade systemic inflammation and an increased incidence of autoimmune diseases [21–23], thus they may further contribute to immune dysregulation in FND patients. However, to date the influence of these factors on the relationship between immune function and FND has not been examined.

To begin addressing this knowledge gap, we conducted a retrospective study to assess the prevalence of several common autoimmune diseases (ADs) in a large cohort of patients with FND and to determine whether this association differed according to biological sex and psychiatric comorbidity. We hypothesized that the prevalence of ADs in patients with FND would be higher than that reported in the general population, with female patients and those with psychiatric comorbidities showing greater rates of ADs.

## 2. Materials and Methods

We conducted a retrospective cohort study using demographic and clinical data collected in the Mass General Brigham (MGB) Biobank. The MGB Biobank is a large biorepository that recruits patients throughout hospitals and clinics across the MGB system [24,25]. All adult patients able to provide informed consent are eligible to participate in this ongoing research project. Consent is provided when patients sign a Data and Sample Use Agreement, which allows MGB researchers to utilize their de-identified health records. The MGB Biobank contains biospecimens and data linked to extensive electronic health record data and survey data of over 135,000 participants. The Mass General Brigham Institutional Review board approved the study.

### 2.1 FND and clinical data

Using the MGB Biobank Portal Query tool, we compiled a cohort of patients 18 years of age or older who received a diagnosis of FND at the Brigham and Women’s Hospital and/or at the Massachusetts General Hospital between January 2015 and December 2021. FND diagnosis was determined by the presence of at least one ICD-10 (International Classification of Diseases, 10th edition) code in a patient’s electronic health record. Specifically, the following ICD-10 codes were used to search for distinct FND subtypes: F44.4 [‘Dissociative motor disorders,’ ‘Conversion disorder with motor symptom or deficit,’ ‘Dissociative motor disorders,’ ‘Psychogenic aphonia,’ ‘Psychogenic dysphonia’] and F44.5 [‘Conversion disorder with seizures or convulsions,’ ‘Dissociative convulsions’]. We further extracted demographic and baseline data (age, sex, race/ethnicity, smoking status, body mass index [BMI]) and medication history), and ascertained the presence of comorbid autoimmune diseases (ADs), neurological disorders (epilepsy and movement disorders), and psychiatric illnesses, using the ICD-10 codes of interest. We considered six major groups of ADs (diseases of endocrine, nervous, digestive, and skin system, connective tissue disorders, and other ADs) [Supplementary Table1]. Patients with comorbid bipolar disorder, schizophrenia, and other psychotic disorders, as assessed by the respective ICD-10 codes, were not included in the study cohort.

### 2.2 Statistical Analysis

We calculated the proportion of FND patients with and without diagnosis of ADs, and the corresponding 95% confidence intervals (CI). In addition to a diagnosis of any ADs, we also examined the prevalence of each of the six groups of ADs as well as the proportion of individuals with multiple disorders (i.e., ≥2 ADs). We further performed subgroup analysis by specific psychiatric comorbid diagnoses (PTSD and/or MDD). Analyses were conducted both across and within sexes and chi-square tests were used to test differences in rates of ADs diagnosis by sex. Odds ratio and 95% confidence intervals were calculated to assess the odds of ADs to occur in women vs men and in subjects with and without psychiatric comorbidities. Differences in demographics and clinical characteristics between subgroups of interest were assessed using chi square test for binary variables and the Student’s t-test for continuous variables. All statistical analyses were performed using GraphPad Prism software version 8.0, with a significance level of *p*□<□0.05.

## 3. Results

In our cohort of 643 individuals with FND, the majority were female (76.3%; *n*□=□491) and identified as Caucasian (80.7%; *n*□=□519). The average age was 48.9 years (± 16.7). Of the 643 participants, 366 (56.9%) had a diagnosis of functional movement disorder [FMD], 201 (31.2%) were diagnosed with functional seizures [FS], while the remaining subjects were diagnosed with both FS and FMD (11.8%; *n*□=□76). Approximately 28% of patients with FND also had a concurrent diagnosis of other neurological disorders (n=182; 138 females and 44 males).

The prevalence of MDD and/or PTSD in our sample was elevated (71.5%; *n*□=□460; 359 females and 101 males), with 178 individuals (38.7%) having a concurrent diagnosis of MDD [single episode or recurrent], 139 (30.2%) diagnosed with PTSD, and 143 meeting criteria for both MDD and PTSD (31%). Of the 183 subjects without psychiatric comorbidities, 134 were females and 49 were males.

### 3.1 Prevalence of ADs

We identified 270 individuals (41.9% [95% CI: 38.1%-45.9%]) with a comorbid diagnosis of ADs among the patients with FND included in our study. The mean age of subjects with ADs was greater than in those without ADs (*t* = 3.0185; *p*= 0.002), whereas no group-differences in race, BMI and smoking status were found (*p*> 0.05) [Table 1]. Among the 6 major groups of ADs considered, connective tissue disorders (i.e., rheumatoid arthritis, systemic lupus, scleroderma) were the most prevalent [33.3 %; *n*□=□90], followed by endocrine ADs (i.e., type I diabetes, Hashimoto’s thyroiditis, Graves’ disease) [31.1 %; *n*□=□84]. Furthermore, of 270 participants with ADs, 75 subjects had ≥2 ADs (27.7% [95% CI: 22.5%-33.5%]) and 24 (8.0% [95% CI: 5.0%-12.9%]) met criteria for multiple autoimmune syndromes (≥3 ADs).

**Table 1.** Distribution and comparisons of demographic and clinical variables among FND patients with and without ADs.

|  | Patients with<br>ADs (n=270) | Patients without ADs<br>(n=373) | *p* value |
| --- | --- | --- | --- |
| Age | 51.3 ± 16.7 | 47.3 ± 16.5 | 0.002 |
| Sex, F/M | 221/49 | 270/103 | 0.005 |
| Caucasian | 221 (81.9%) | 299 (80.2%) | 0.59 |
| BMI | 30.3 ± 7.72 | 29.7 ± 7.31 | 0.31 |
| Smokers | 8 (2.96%) | 13 (3.49%) | 0.71 |
| Neurological comorbidities | 80 (29.6%) | 102 (27.3%) | 0.4 |

Results of a chi-square test indicated that the prevalence of ADs significantly differed among sexes (*X*^2^□=□7.77, *p*□=□0.005) [Table 1], with women showing a prevalence of 45.0% (n□=□221/491; [95% CI: 40.5%-49.5%]), compared to 32.2% in men (n□=□49/152; [95% CI: 24.8%-40.2%]). The odds of having ADs among female patients with FND were 1.72 (95% CI: 1.17-2.53) times the odds of that of male patients with FND. Female patients with FND and ADs were significantly younger compared to male patients (*p*□=□0.0001), while no sex-differences in other demographic and clinical variables were found (*data not shown*).

Next, we assessed whether the prevalence of ADs in patients with FND varied as a function of psychiatric comorbidity (MDD and/or PTSD). In the subgroup of FND patients with comorbid MDD and/or PTSD, 44.7% of the subjects also had a diagnosis of ADs [206/460; 95% CI: 40.1%-49.4%] compared to 34.9% [64/183; 95% CI: 28.0% - 42.3%] in patients without psychiatric comorbidities (*X*^2^□=□5.17; *p*□=□0.02; OR= 1.5; 95 % CI:1.05-2.15). Women represented the majority of individuals with ADs both in the subgroup with comorbid MDD and/or PTSD (78.1%; 95% CI: 76.6%-87.4%) as well as among subjects without psychiatric comorbidities (72.1%; 95% CI: 67.7%-88.7%) (*X*^2^□=□0.26; *p*□=□0.60)

## 4. Discussion

To our knowledge, this is the first study to assess the prevalence of ADs in a large sample of men and women with FND, and to evaluate the role of psychiatric comorbidities and biological sex in modulating this association. The overall prevalence of ADs in our sample was 41.9%, while rates of 10% have been reported in the general population (13% in women and 7% in men), according to a recent retrospective population-based study [26]. Connective tissue disorders and autoimmune endocrine diseases were the most common ADs observed in our FND sample. We further found that the prevalence of ADs was significantly higher in female patients with FND and in those with co-occurring MDD and/or PTSD. However, prevalence rates of ADs were greater than those observed in the general population even in absence of psychiatric comorbidities, thus supporting a link between FND and immune dysfunction.

These findings are in line with previous studies reporting that compared to healthy controls, patients with FND have elevated levels of pro-inflammatory molecules, including IL-1, IL-6, IFN-γ, and TNF-α [16,27], and increased levels of immunoglobulins [12] and C-reactive protein (CRP) [13]. These observations suggest that FND may be characterized by a low-grade inflammatory state, which could serve as one molecular mechanism underlying several brain and behavioral alterations associated to this disorder. Indeed, higher levels of endogenous inflammatory markers have been linked to abnormalities in functional connectivity in neurocircuits and networks involved in motivation and motor activity, threat detection, anxiety, and interoceptive and emotional processing (for a review see [28]). At a clinical level, low-grade inflammation has been implicated in the development of chronic pain and fatigue [29,30], and in the emergence of mood disturbances, which are frequently reported by patients with FND. Epidemiological studies further support the hypothesis of an association between FND and inflammation by showing that traumatic injuries or exposure to other physical and/or psychological stressors – which activate inflammatory response in brain as well as peripherally [31,32]–often precede the onset or worsening of functional neurological symptoms [33–35]. Interestingly, stress exposure is also involved in triggering and exacerbating ADs [23,36], thus representing a potential mechanism linking the emergence of both functional neurological symptoms and alterations in immune response. To better characterize this relationship, it will be critical to determine whether the relationship between FND and ADs is bidirectional, and whether changes in inflammatory response precedes or follows the development of FND.

We further observed that the proportion of women diagnosed with ADs in our cohort was significantly higher than men, both in the overall sample and in the subgroups of interest (patients with and without comorbid MDD and/or PTSD), mirroring the gender-bias in the prevalence of ADs observed in the general population [37]. This bias likely reflects sex-differences in immune response, which is influenced by a complex interplay between sex hormones, sex chromosomes, immune-related genes and environmental exposure [18]. As a result, females exhibit a more significant immune response and therefore increased vulnerability towards ADs. Evidence that immunopathogenic mechanisms are also implicated in FND may provide critical insights into the biological underpinnings of the well-documented but often poorly explained predominant prevalence of FND among females [38,39]. As such, future studies investigating inflammatory processes in patients with FND should specifically consider the role of biological sex.

We also found that the association between FND and ADs was stronger in presence of psychiatric comorbidities (MDD and/or PTSD). Indeed, patients affected by these psychiatric disorders exhibit abnormal levels of cytokines [21,22,40] and have an increased vulnerability for ADs [23]. Thus, it is possible that shared risk factors, particularly environmental stressors, may induce a common core of molecular alterations underlying FND, stress-related psychiatric disorders and ADs. However, in subjects without psychiatric comorbidities, the prevalence of ADs was still significantly greater than that observed in the general population. As such, the link between immune dysregulation and FND may be independent of, yet probably amplified by, the presence of psychiatric comorbidities. A compelling question is whether specific alterations in immune response and/or inflammatory markers profiles are associated with FND compared to other disorders and could therefore represent an FND-specific disease biomarker.

Our findings should be interpreted in light of several limitations. The primary limitation of the current study is its retrospective nature which precludes any causal inferences regarding the association between FND and ADs. Second, our sample consisted of patients attending hospitals, clinics and tertiary care centers, which may explain the high rate of ADs observed. However, individuals with FND often undergo extensive diagnostic tests, which may facilitate the identification of comorbid conditions in this patient population compared to others. Third, while a large sample size is a strength of this study, diagnostic and clinical data were obtained from electronic medical records and not from prospectively delivered structured clinical interviews and exams. We, therefore, cannot determine with certainty the reliability of all diagnoses (or absence thereof), although medication history was used as a proxy to confirm diagnosis of ADs, MDD and PTSD. Furthermore, we lacked measures of severity or specific phenomenology within FND subtypes, as well as data regarding exposure to childhood trauma, which should be considered in future research. These limitations are inherent to retrospective cohort studies using electronic health records, which, however, offer an extremely valuable opportunity to explore novel research questions using larger sample size and could help inform prospective studies [41,42].

In summary, we identified a high prevalence of ADs in a cohort of 643 patients with FND and demonstrated that female sex and psychiatric comorbidities increase the strength of this association. These findings further support prior studies showing alterations in inflammatory response and immune-related pathways in FND and have relevance both in terms of understanding the underlying disease pathophysiology as well as for the clinical management of patients with FND. Future studies are needed to better understand the association between ADs and FND and to identify FND-specific immune dysregulations.

## Contributorship

PAS, GB and BAD conceived the original idea. AJ and PAS wrote the initial draft of this manuscript. AJ, GB, and ANC performed data collection and processing. AJ and PAS performed data analysis with help from GB and ANC. GRP, ANC, IG and MAO contributed to the interpretation of the results. PAS supervised the project. All authors provided critical feedback and helped shape the research, analysis and manuscript.

## Data Availability

All data produced in the present study are available upon reasonable request to the authors

## Acknowledgements

This work has been supported by the Mary Ann Tynan Fellowship Program and the Women’s Brain Initiative/Brigham and Women’s Hospital. We also acknowledge the participants and administrators of the Mass General Brigham Biobank for their contribution to this work.

## References

1 Hallett M, Aybek S, Dworetzky BA, et al. Functional neurological disorder: new subtypes and shared mechanisms. Lancet Neurol. 2022;21:537–50.

2 Spagnolo PA, Garvey M, Hallett M. A dimensional approach to functional movement disorders: Heresy or opportunity. Neurosci Biobehav Rev. 2021;127:25–36.

3 Patron VG, Rustomji Y, Yip C, et al. Psychiatric Comorbidities in Functional Neurologic Symptom Disorder. Pract Neurol (Fort Wash Pa). 2022;21:71–5.

4 Erro R, Brigo F, Trinka E, et al. Psychogenic nonepileptic seizures and movement disorders. Neurol Clin Pract. 2016;6:138–49. doi: 10.1212/CPJ.0000000000000235

5 Wissel BD, Dwivedi AK, Merola A, et al. Functional neurological disorders in Parkinson disease. J Neurol Neurosurg Psychiatry. 2018;89:566–71. doi: 10.1136/jnnp-2017-317378

6 Stone J, Carson A, Duncan R, et al. Who is referred to neurology clinics?--the diagnoses made in 3781 new patients. Clin Neurol Neurosurg. 2010;112:747–51.

7 Espay AJ, Aybek S, Carson A, et al. Current Concepts in Diagnosis and Treatment of Functional Neurological Disorders. JAMA Neurol. 2018;75:1132–41.

8 Paredes-Echeverri S, Maggio J, Bègue I, et al. Autonomic, Endocrine, and Inflammation Profiles in Functional Neurological Disorder: A Systematic Review and Meta-Analysis. JNP. 2022;34:30–43.

9 Aybek S, Perez DL. Diagnosis and management of functional neurological disorder. BMJ. 2022;376:o64.

10 Maurer CW, LaFaver K, Ameli R, et al. Impaired self-agency in functional movement disorders: A resting-state fMRI study. Neurology. 2016;87:564–70.

11 Winterdahl M, Miani A, Vercoe MJH, et al. Vulnerability to psychogenic non-epileptic seizures is linked to low neuropeptide Y levels. Stress. 2017;20:589–97.

12 Khanam M, Ullah MdA, Shamsul M, et al. Serum Immunoglobulin Profiles of Conversion Disorder Patients. German Journal of Psychiatry. 2008;11.

13 Kozlowska K, Chung J, Cruickshank B, et al. Blood CRP levels are elevated in children and adolescents with functional neurological symptom disorder. Eur Child Adolesc Psychiatry. 2019;28:491–504.

14 Tiyekli U, Çalıyurt O, Tiyekli ND. Proinflammatory cytokine levels in patients with conversion disorder. Acta Neuropsychiatrica. 2013;25:137–43.

15 Nagata R, Matsuura E, Nozuma S, et al. Anti-ganglionic acetylcholine receptor antibodies in functional neurological symptom disorder/conversion disorder. Front Neurol. 2023;14:1137958.

16 van der Feltz-Cornelis C, Brabyn S, Ratcliff J, et al. Assessment of cytokines, microRNA and patient related outcome measures in conversion disorder/functional neurological disorder (CD/FND): The CANDO clinical feasibility study. Brain Behav Immun Health. 2021;13:100228.

17 Spagnolo PA, Johnson K, Hodgkinson C, et al. Methylome changes associated with functional movement/conversion disorder: Influence of biological sex and childhood abuse exposure. Prog Neuropsychopharmacol Biol Psychiatry. 2023;125:110756.

18 Klein SL, Flanagan KL. Sex differences in immune responses. Nat Rev Immunol. 2016;16:626–38.

19. Rubinow DR, Schmidt PJ. Sex differences and the neurobiology of affective disorders. Neuropsychopharmacol. 2019;44:111–28.

20 Tolin DF, Foa EB. Sex differences in trauma and posttraumatic stress disorder: A quantitative review of 25 years of research. Psychological Bulletin. 2006;132:959–92.

21 Powers A, Dixon HD, Conneely K, et al. The differential effects of PTSD, MDD, and dissociation on CRP in trauma-exposed women. Compr Psychiatry. 2019;93:33–40.

22 Renner V, Schellong J, Bornstein S, et al. Stress-induced pro- and anti-inflammatory cytokine concentrations in female PTSD and depressive patients. Transl Psychiatry. 2022;12:1–6. doi: 10.1038/s41398-022-01921-1

23 Song H, Fang F, Tomasson G, et al. Association of Stress-Related Disorders With Subsequent Autoimmune Disease. JAMA. 2018;319:2388–400.

24 Castro VM, Gainer V, Wattanasin N, et al. The Mass General Brigham Biobank Portal: an i2b2-based data repository linking disparate and high-dimensional patient data to support multimodal analytics. Journal of the American Medical Informatics Association. 2022;29:643– 51.

25 Boutin NT, Schecter SB, Perez EF, et al. The Evolution of a Large Biobank at Mass General Brigham. J Pers Med. 2022;12:1323.

26 Conrad N, Misra S, Verbakel JY, et al. Incidence, prevalence, and co-occurrence of autoimmune disorders over time and by age, sex, and socioeconomic status: a population-based cohort study of 22 million individuals in the UK. The Lancet. 2023;401:1878–90.

27 Tiyekli U, Calıyurt O, Tiyekli ND. Proinflammatory cytokine levels in patients with conversion disorder. Acta Neuropsychiatr. 2013;25:137–43.

28 Goldsmith DR, Bekhbat M, Mehta ND, et al. Inflammation-Related Functional and Structural Dysconnectivity as a Pathway to Psychopathology. Biological Psychiatry. 2023;93:405–18.

29 Parkitny L, McAuley JH, Di Pietro F, et al. Inflammation in complex regional pain syndrome: a systematic review and meta-analysis. Neurology. 2013;80:106–17.

30 Raison CL, Rye DB, Woolwine BJ, et al. Chronic interferon-alpha administration disrupts sleep continuity and depth in patients with hepatitis C: association with fatigue, motor slowing, and increased evening cortisol. Biol Psychiatry. 2010;68:942–9.

31. Rohleder N. Stimulation of Systemic Low-Grade Inflammation by Psychosocial Stress. Psychosomatic Medicine. 2014;76:181–9.

32 Calcia MA, Bonsall DR, Bloomfield PS, et al. Stress and neuroinflammation: a systematic review of the effects of stress on microglia and the implications for mental illness. Psychopharmacology. 2016;233:1637–50.

33 Ludwig L, Pasman JA, Nicholson T, et al. Stressful life events and maltreatment in conversion (functional neurological) disorder: systematic review and meta-analysis of case-control studies. Lancet Psychiatry. 2018;5:307–20.

34 Jobin K, Wang M, du Plessis S, et al. The importance of screening for functional neurological disorders in patients with persistent post-concussion symptoms. NeuroRehabilitation. 2023;53:199–208.

35 Stone J, Carson A, Aditya H, et al. The role of physical injury in motor and sensory conversion symptoms: A systematic and narrative review. Journal of Psychosomatic Research. 2009;66:383–90.

36 Keynejad RC, Frodl T, Kanaan R, et al. Stress and functional neurological disorders: mechanistic insights. J Neurol Neurosurg Psychiatry. 2019;90:813–21.

37 Angum F, Khan T, Kaler J, et al. The Prevalence of Autoimmune Disorders in Women: A Narrative Review. Cureus. 2020;12:e8094.

38 Baizabal-Carvallo JF, Jankovic J. Gender Differences in Functional Movement Disorders. Mov Disord Clin Pract. 2020;7:182–7.

39 Lidstone SC, Costa-Parke M, Robinson EJ, et al. Functional movement disorder gender, age and phenotype study: a systematic review and individual patient meta-analysis of 4905 cases. J Neurol Neurosurg Psychiatry. 2022;93:609–16.

40 Euesden J, Danese A, Lewis CM, et al. A bidirectional relationship between depression and the autoimmune disorders – New perspectives from the National Child Development Study. PLoS One. 2017;12:e0173015.

41 Sauer CM, Chen L-C, Hyland SL, et al. Leveraging electronic health records for data science: common pitfalls and how to avoid them. The Lancet Digital Health. 2022;4:e893–8.

42 Dziadkowiec O, Durbin J, Jayaraman Muralidharan V, et al. Improving the Quality and Design of Retrospective Clinical Outcome Studies that Utilize Electronic Health Records. HCA Healthcare Journal of Medicine. 2020;1. doi: 10.36518/2689-0216.1094

