## Supplemental Table 1 for "Prevalence of Autoimmune Diseases in Functional Neurological Disorder: Influence of Psychiatric Comorbidities and Biological Sex"

Supplemental Table 1. Diseases and their respective ICD-10 codes used to identify patients with FND included in the study cohort

| ***Diseases*** |  | ***ICD-10 codes*** |
| --- | --- | --- |
| *Autoimmune Diseases* | | |
| **Disease of endocrine system** | Diabetes mellitus, insulin dependent | E10 |
|  | Autoimmune thyroid disease | E03.5, E03.9, E05.0, E05.5, E05.9, E06.3, E06.5 |
|  | Addison’s disease | E27.1, E27.2 |
| **Connective tissue disorders** | Systemic lupus erythematosus | M32 |
|  | Systematic sclerosis (scleroderma) | M34 |
|  | Sjögren’s syndrome | M35.0 |
|  | Rheumatoid arthritis | M08.1, M08.2, M08.3, M08.4 |
| **Disease of skin system** | Dermatitis herpetiformis | L13.0 |
|  | Psoriasis | L40 |
|  | Alopecia areata | L64 |
| **Disease of the digestive system** | Primary biliary cirrhosis | K74.3 |
|  | Coeliac disease | K90.0 |
|  | Crohn’s disease | K50 |
|  | Ulcerative colitis | K51 |
| **Disease of nervous system** | Multiple sclerosis | G35 |
|  | Guillain-Barré syndrome | G61.9 |
|  | Myasthenia gravis | G70.0 |
| **Other AIDs** | Immune thrombocytopenic purpura | D69.3 |
|  | Sarcoidosis | D86 |
| *Psychiatric Disorders* | | |
| **Major Depressive Disorder** | Single Episode | F32.9 |
|  | Recurrent | F33 |
| **PTSD** |  | F43.1 |
| **Schizophrenia, schizotypal, delusional, and other non-mood psychotic disorders** |  | F20-29 |
| **Bipolar Disorder** |  | F31 |
| *Neurological Disorders* | | |
| **Movement Disorders** |  | G20-G26 |
| **Epilepsy** |  | G40 |
